# SARS-CoV-2 Control on a Large Urban College Campus Without Mass Testing

**DOI:** 10.1101/2021.01.21.21249825

**Authors:** Chris O’Donnell, Kate Brownlee, Elise Martin, Joe Suyama, Steve Albert, Steve Anderson, Sai Bhatte, Kenyon Bonner, Chad Burton, Micaela Corn, Heather Eng, Bethany Flage, Jay Frerotte, Goundappa K. Balasubramani, Catherine Haggerty, Joel Haight, Lee H. Harrison, Amy Hartman, Thomas Hitter, Wendy C. King, Kate Ledger, Jane W. Marsh, Margaret C. McDonald, Bethany Miga, Kim Moses, Anne Newman, Meg Ringler, Mark Roberts, Terrie Sax, Anantha Shekhar, Matthew Sterne, Tyler Tenney, Marian Vanek, Alan Wells, Sally Wenzel, John Williams

## Abstract

**Objective:** A small percentage of universities and colleges conduct mass SARS-CoV-2 testing. However, universal testing is resource-intensive, strains national testing capacity, and false negative tests can encourage unsafe behaviors.

**Participants:** A large urban university campus.

**Methods:** Virus control centered on three pillars: mitigation, containment, and communication, with testing of symptomatic and a random subset of asymptomatic students.

**Results:** Random surveillance testing demonstrated a prevalence among asymptomatic students of 0.4% throughout the term. There were two surges in cases that were contained by enhanced mitigation and communication combined with targeted testing. Cumulative cases totaled 445 for the term, most resulting from unsafe undergraduate student behavior and among students living off-campus. A case rate of 232/10,000 undergraduates equaled or surpassed several peer institutions that conducted mass testing.

**Conclusions:** An emphasis on behavioral mitigation and communication can control virus transmission on a large urban campus combined with a limited and targeted testing strategy.

## INTRODUCTION

The ongoing SARS-CoV-2 pandemic has major implications for institutions of higher education (IHE). Approaches for monitoring student infections vary among institutions, with some conducting minimal or no testing while others opt for mass testing ^1^. Here, we present the University of Pittsburgh’s (Pitt) multidisciplinary approach adopted during the fall 2020 term. The foundation of virus control on Pitt’s main campus had three pillars: 1) mitigation; 2) communication; and 3) containment (symptomatic and surveillance testing, contact tracing, and isolation/quarantine)—and was unique in its strong reliance on mitigation over mass testing.

### Framework and policies

Pitt is a public research university with five campuses. The focus of this paper is on the largest campus, the Pittsburgh campus. In the 2020 fall term, the Pittsburgh campus had 28,234 students, including 19,197 undergraduate students and 13,264 faculty and staff members. The Pittsburgh campus is located in the Oakland neighborhood, an urban area of Pittsburgh and part of Allegheny County, home to a population of 1.2 million. Pitt devised a framework of operational Risk Postures (Guarded, Elevated, and High), which informed decision making and the level of on-campus activities, and Flex@Pitt, an instructional model that accommodated both in-person and remote instruction. A Health Care Advisory Group (HCAG) chaired by the Dean of the School of Medicine included experts in epidemiology, general internal medicine, disease and risk-modeling, environmental and occupational health, pulmonology, infectious disease, health and risk communication, law, and policy. The HCAG developed campus-wide standards and guidelines on the use of masks, personal protective equipment, shared spaces, travel, and other pertinent issues. The HCAG assessed Risk Posture weekly (and as needed) based on a comprehensive assessment of multiple parameters, including infection rate, isolation capacity, hospital capacity, testing and contract tracing capacity, local and national incidence rates, and others. An Implementation and Oversight Committee (IOC) was formed with subcommittees led by virus testing experts, behavioral experts, and students. The COVID-19 Medical Response Office (CMRO) comprised faculty trained in infectious diseases, infection prevention, and pandemic preparedness, and was charged with operationalizing the day-to-day management of virus control on the campus. All committees met at least weekly and ideas and information flowed freely between groups. The CMRO was integrated into committees and groups central to managing the pandemic. Members of Pitt Athletics were tested per National Collegiate Athletic Association guidelines by an outside vendor and are not further discussed herein, but positive student athlete results were included in the totals presented in this work. The Resilience Steering Committee, chaired by the Senior Vice Chancellor and Chief Legal Officer, brought together key personnel and student leaders to coordinate, operationalize and disseminate information.

### Semester schedule

The start and end dates of the semester were changed so that classes began remotely 8/19, with 11/20 the last day of classes. In-person final examinations occurred 11/23-24, with Thanksgiving recess 11/25-29 and remote-only finals 11/30-12/3. Thus, students did not return after Thanksgiving break.

### Housing

A multidisciplinary team consisting of Housing, Dining, Student Affairs, Student Health Services (SHS), Environmental Health & Safety (EHS), Infectious Disease, Epidemiology, and Public Health members developed housing guidelines with an aim to meet or exceed recommendations from the Centers for Disease Control and Prevention (CDC) and local/state health departments regarding physical distancing, airflow, and sanitation. Approximately one-third of Pitt undergraduates (∼6300 in fall 2020) lived in on-campus university student housing or university-rented hotels. Configurations of university housing include high rise buildings with communal bathrooms, individual and shared rooms, suites (multi-bedroom apartments without kitchens), and apartments (units with kitchens). To mitigate risk during the pandemic, Pitt de-densified double rooms to singles and units for 3-4 students were limited to 2 students on floors with communal bathrooms. Pitt augmented its housing capacity by contracting extra rooms with local hotels. Floors/wings with communal bathrooms were limited to 31 students and communal bathroom use was limited to <10 students at one time. Housekeeping was enhanced for all communal bathrooms using EPA-registered disinfectants at least twice per day. Pitt provided supplies for residents to clean surfaces between staff cleanings. Signage instructed students on proper handwashing and techniques to minimize contact with bathroom surfaces. Residential suites, apartments, and hotel rooms with shared bathrooms were housed at capacities of between 2-8 students each. Doors and some sinks were refitted for hands-free operation.

To mitigate risk during student arrivals at the start of the academic year, Pitt implemented a staggered campus repopulation plan. This plan used a shelter-in-place model with pods of 4-6 students acting as a functional household of close contacts and accommodated five student cohorts arriving over 2 days each, every fourth day during a 2-week period at the beginning of fall term. Move-in procedures and schedules were extensively modified—including limiting each incoming student to one personal contact to provide move-in support—to maintain physical distancing and minimize close contacts.

### Facilities

Pitt conducted a University-wide assessment of HVAC systems. Independent ventilation experts partnered with internal personnel to maximize outdoor supply of air and assure adequate air change rates in all buildings and rooms. A few rooms were deemed inadequate due to a high percentage of indoor air re-circulation or a low rate of air changes per hour. Maximum occupancy limits were posted for all shared spaces, including classrooms, meeting rooms, break rooms, and laboratories. Most assessments were performed using floor plans but numerous sites were visited to verify space configuration.

### Student involvement

Pitt deemed student involvement essential to encourage effective mitigation behaviors. The student members of the IOC developed a sub-committee composed only of students, which served as a forum for questions, ideas, feedback, and new initiatives. The sub-committee encompassed students from different educational disciplines, social groups, and educational levels, and included both leaders and non-leaders with a ratio of graduates to undergraduates that roughly reflected the student body. This group met virtually each week throughout the term, encouraged honest feedback, and fostered peer-to-peer conversations. Detailed notes were shared with the Dean of Students and the IOC, including student experiences and campus rumors. This real-time feedback informed changes in guidelines, policies, and communications. Direct, bidirectional, and iterative communication with students occurred through multiple other channels, including campus-wide meetings as well as meetings with targeted student groups, such as Greek organizations, student government, residential assistants, student safety ambassadors, and student clubs.

### Culture

A diverse committee of undergraduate and graduate students wrote the Pitt Community Compact, a commitment by members of the University community to embrace a culture of behaviors that supported the safety and well-being of self and others. Pitt hired Off-Campus Student Safety Ambassadors, a team of >30 students who conducted rounds in pairs throughout the campus and adjacent neighborhoods and conversed with students while distributing public health materials and information. Other culture-focused strategies included nighttime safety walks by the Dean of Students through neighborhoods adjacent to campus, where students were most likely to reside in off-campus residences. These walks were designed to engage and educate students, connect with neighborhood residents and reinforce positive behaviors in relation to mask wearing and physical distancing. During these walks, administrators also provided masks to those who needed them as well as Pitt merchandise to students following rules.

### Communication

Pitt implemented regular text messages, emails, videos, and social media posts exhorting the use of masks, physical distancing, and proper hand hygiene. These messages amplified residential policies that enjoined students to remove masks only within their own pod. Local businesses, restaurants and bars also supported distancing efforts and curfew. The Office of University Communications and Marketing (UCM) and Community and Governmental Relations worked with the Student Government Board and Oakland Business Improvement District to provide signage to area businesses promoting healthy behaviors, to reinforce behaviors on and off campus. The goal was to get students to consider *community protection*, since it was recognized that students would (correctly) conclude that they were at low risk for severe COVID-19. Ongoing messaging— including a targeted marketing campaign, the Power of Pitt—reinforced the idea that an individual’s behavior could indirectly harm others, including vulnerable community members, as well as threaten the student body’s ability to safely remain on campus (**Figure 1**). Additionally, weekly updates were provided to schools and departments across campus with shareable resources, key messages, and upcoming priorities. This ensured consistent messaging was repeated throughout the University, and the most important messages were being amplified.

**Figure 1.**
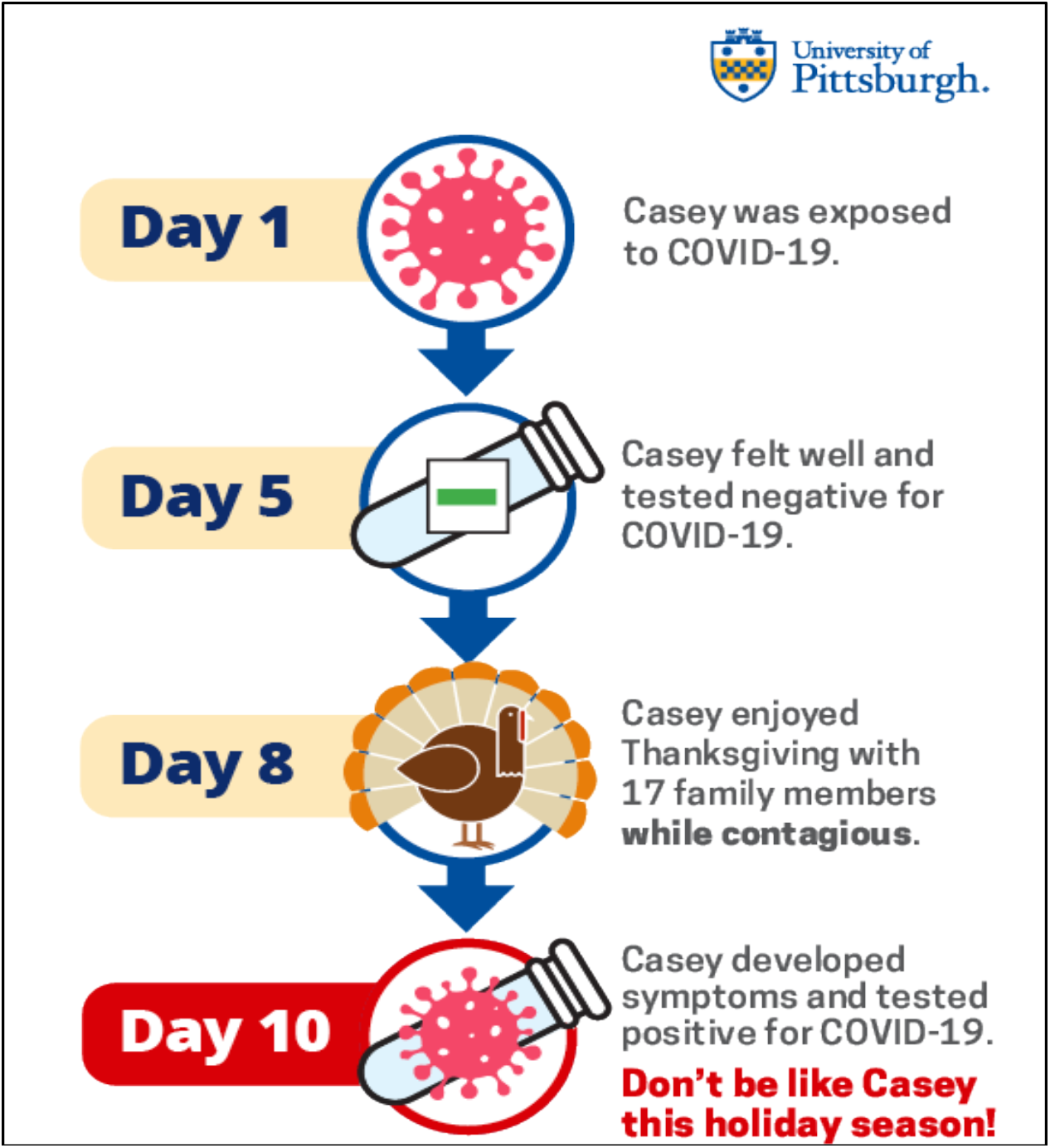
Graphic to illustrate limitations of a single negative test to encourage student mitigation behaviors.

### Coronavirus.pitt.edu

Providing a central source of reliable, frequently updated information was recognized as a need early in the pandemic. <u>Coronavirus.pitt.edu</u> became the hub where University members, community members, parents and others could learn about the University’s response, stay up-to-date on the latest health and safety guidance, report concerns, and read the latest CMRO messages.

### Dashboards

The CMRO developed a public Internet dashboard as well as an internal dashboard with >100 variables, many automatically updated daily. The public dashboard reported the number of positive student and employee tests daily for the prior four weeks; the five-day rolling average; the number of cases in isolation; the cumulative number of positive tests; and data from asymptomatic surveillance testing for the entire term. The public dashboard was updated every Tuesday and Friday. The CMRO disseminated these data with a narrative that included helpful context, reinforced good behavior, praised success, and set expectations when interventions were required. The comprehensive internal dashboard, available to University leadership, offered detailed campus and community data pertaining to the pandemic and the pandemic response capacity locally. University leaders consulted this dashboard before making decisions about changing the institution’s Risk Posture.

### Shelter-in-place

At the beginning of the term, the CMRO asked all students to shelter in place for 14 days, including a minimum period of 7 days after arriving on campus. Behavioral experts indicated that students were unlikely to follow a strict 14-day quarantine period. Consequently, Pitt opted for a less restrictive shelter-in-place strategy that allowed for responsible outdoor dining and activities as well as close contact with a pod of up to 4-6 students. Near the end of the term, the CMRO asked students to complete a second 14-day shelter-in-place period. This shelter-in-place order was designed to help students minimize the risk of transmission as they traveled and arrived home for winter break; students with vulnerable household members at home were counseled to quarantine even from podmates. Most students left campus on or before Nov. 20 after completing a minimum of 10 days shelter in place, with another minimum of four days recommended upon arrival at home.

### Testing

SARS-CoV-2 PCR testing was an integral component of the strategy to monitor incidence, detect cases, and monitor and interrupt transmission. All symptomatic students and some asymptomatic household contacts of cases were tested individually by SHS, the student-serving medical clinic on campus, in a Clinical Laboratory Improvement Amendments (CLIA)-certified laboratory throughout the term. The CMRO invited a random sample of asymptomatic students for surveillance testing during the staged move-in period and throughout the term. Students who were selected but were experiencing symptoms were removed from the asymptomatic sample and sent to SHS for testing. Given evidence that SARS-CoV-2 infection prevalence varies by region, and that spread differs in congregate versus non-congregate settings ^2-4^, the CMRO considered student subgroups defined by where they were coming from: 1) residents of the Western Pennsylvania region; 2) in-state students from outside of Western Pennsylvania; 3) out-of-state domestic students from other US states; and 4) international students. Each group was further divided into on-campus and off-campus residents, resulting in 8 groups overall. The sample size for each group was estimated based on the number of students in a group, with an assumed prevalence of ≤0.75% based on unpublished local serologic data, a desired precision of □1% and 80% power. Sample size and power calculations were performed with PASS version 13.0.1 (NCSS LLC, Kaysville, Utah USA). The calculated subgroup sample sizes ranged from 211 to 274 each for a total of 2055 students.

Before classes began, Pitt established a large central outdoor site for observed self-collected nasal specimens and oversaw the testing of about 10% of the student population during move-in. During campus repopulation (8/12-8/29), specimens were collected within 48 hours of a student’s arrival to campus and results were provided within 24 hours. A large central outdoor site was created for observed self-collected nasal specimens.

Self-collection was observed and supervised using in-person or virtual methods (HIPAA-compliant video). Specimens were barcoded and all data stored in a dedicated REDCap database ^5, 6^, which facilitated automated result notifications and reports. To accommodate CLIA-certified testing, Pitt used an epMotion 5075 pipetting robot (Eppendorf) to achieve 4:1 pooling; this ratio was chosen to limit the loss of analytical sensitivity to a cycle threshold (Ct) change of 2 and was validated as a laboratory developed test (LDT) as defined by the National Committee for Clinical Laboratory Standards. The four individual specimens from a positive pool were tested to identify which student(s) were SARS-CoV-2 positive, using either the Hologic Panther or an LDT based on the CDC Emergency Use Authorization (EUA) protocol. Both platforms had EUA for clinical testing and were validated for pooling.

For the ongoing surveillance effort over the fall term, calculations were performed to determine the magnitude of change in prevalence that could be detected with a sample size of ∼500 per week assuming random selection of asymptomatic students with no replacement. Based on the prevalence of positive asymptomatic students during the move-in period, a change in prevalence from 0.31% to 2.33%% in a single week could be detected with 80% power and 0.05 significance level. An upward trend over multiple weeks could be detected with smaller increases per week.

Ongoing surveillance testing after move-in continued using pooling, but testing was performed in a university laboratory using the same CDC protocol (and cross validated to the CLIA-certified assay); all positives (and numerous negatives) detected via surveillance testing were confirmed in a secondary assay. Each pool that tested positive during surveillance had at least one individual specimen test positive, validating the method. All collections sent for pooling had a positive control specimen sent to assess pooling quality. Ongoing testing also included “focused testing” for students and staff associated with clusters (e.g., specific residences or groups) or for asymptomatic close contacts of positive students. Upon returning to their permanent residence at the end of the term, the CMRO offered students free optional self-collected testing through Quest laboratories and advised to continue a minimum of 4 days shelter in place. Residual specimens were stored for possible genomic epidemiologic analysis.

### Contact tracing

Pitt performed tracing of all persons testing positive for SARS-CoV-2, including asymptomatic surveillance students. Tracing was managed by SHS and EH&S using a tracing team largely staffed by paid public health graduate students. Tracers maintained confidentiality and did not provide information on the index case. To encourage compliance and honesty, communications to students reiterated that contact tracers did not share data with the Student Conduct Office, and no student would be penalized based on conversations with the contact tracer, even if their behavior violated University rules. Each student who tested positive received an email confirming their 14-day quarantine window and sharing helpful information and resources. Tracers were available seven days per week and most tracing was performed on the same day that exposed individuals were identified. Tracers made multiple attempts by phone and email to contact all exposed individuals. All quarantine and isolation cases were entered into a Pitt-developed COVID-19 tracker tool. These data were used to monitor trends. Data on positive cases were shared with the local county health department, which reciprocated by communicating index case and close contact information for those community-acquired cases known to be Pitt-affiliated.

### Isolation/quarantine

Students who tested positive for SARS-CoV-2 were instructed to isolate for a period of 10 days. They had the option of isolating in designated on-campus housing or at an off-campus location of their choosing (including their permanent residence). If a student opted to isolate off-campus, SHS confirmed that the location was safe and allowed for appropriate physical distancing. If on campus, the COVID Support Team performed daily check-ins and meal delivery. SHS reached out daily to each student in isolation—regardless of their location—to answer their questions and address medical needs, monitor recovery and release them from isolation when it was medically safe to do so. This approach evolved due to student feedback, which inspired the distribution of gift bags at isolation mid-point; resource links to Campus Recreation, SHS, and the Counseling Center; modified meal delivery times; and the addition of laundry services. SHS also augmented its support capacity by hiring part-time nurses and implementing a 24-hour nurse hotline to triage COVID-related inquiries.

Students with known or suspected exposure to SARS-CoV-2 were instructed to quarantine for 14 days. They would only be tested if they developed symptoms while in quarantine. Those quarantining on campus also received daily check-ins and meal delivery from the COVID Support Team. These students could not leave quarantine except to receive medical care, including COVID-19 testing, which Pitt offered to all known exposed students. Students were allowed to quarantine off campus if their residence could accommodate safe physical distancing. If the index case was an off-campus roommate, the exposed student was instructed to avoid all contact with the index case. All students, regardless of their quarantine location, were instructed to log symptoms online daily. University contact tracers could release cases from quarantine at the appropriate time.

### Compliance

Oversight of community compliance was delegated to a COVID-19 Health & Safety Compliance Team, which included representatives from the following Pitt units: Student Affairs, Athletics, Community and Governmental Relations, Housing and Dining, Public Safety, Student Conduct, CMRO, Communications, and the Office of Compliance, Investigations and Ethics. This team reviewed data from the COVID Concern reporting system, student conduct referrals, and university police reports among other sources. The team used these data to inform communication, compliance and testing strategies. Campus compliance with masking and physical distancing was monitored by 40 Concierge Stations on the campus. In addition, structured de-identified observations, were randomly collected at designated campus locations by a team of industrial engineering students. These students performed observation hourly from 08:00 until 23:00 seven days/week, which resulted in ∼140 observations per day and ∼1000 observations per week.

## RESULTS

1906 students during repopulation and 7389 students during the remainder of the fall semester were selected for random asymptomatic surveillance testing. The primary reason for non-participation in the voluntary testing during repopulation was not living near the Pitt campus (i.e., due to remote instruction). The sampling scheme was improved during the fall semester to minimize this problem by selecting students who used a local IP address to login to their Pitt account. In addition to the 7389 students tested via asymptomatic random surveillance, Pitt tested 3102 symptomatic students, 228 close contacts, and 786 students via focused testing, a total of 11,505 students (**Table 1**). Asymptomatic surveillance testing indicated a slight increase during the fall semester following the 18-day arrival period, but it remained low throughout the semester (**Table 1**). The staff/faculty case rate remained low over the summer—despite resuming on-site research in June—and for the duration of the term (**Figure 2**). In September, off-campus socialization fueled a surge in students testing positive for SARS-CoV-2. In response, Pitt launched a communications campaign touting the heightened mitigation efforts and reminding students of possible disciplinary consequences of noncompliance. Pitt also increased focused and close contact testing as well as direct outreach to students including the nighttime neighborhood safety walks described above. Following implementation of these measures, virus levels returned to and remained at baseline levels of <5 student cases/day. As a result, the campus shifted from Elevated to Guarded Posture on Oct. 19, allowing mostly in-person instruction and the resumption of organized student activities.

**Table 1.**
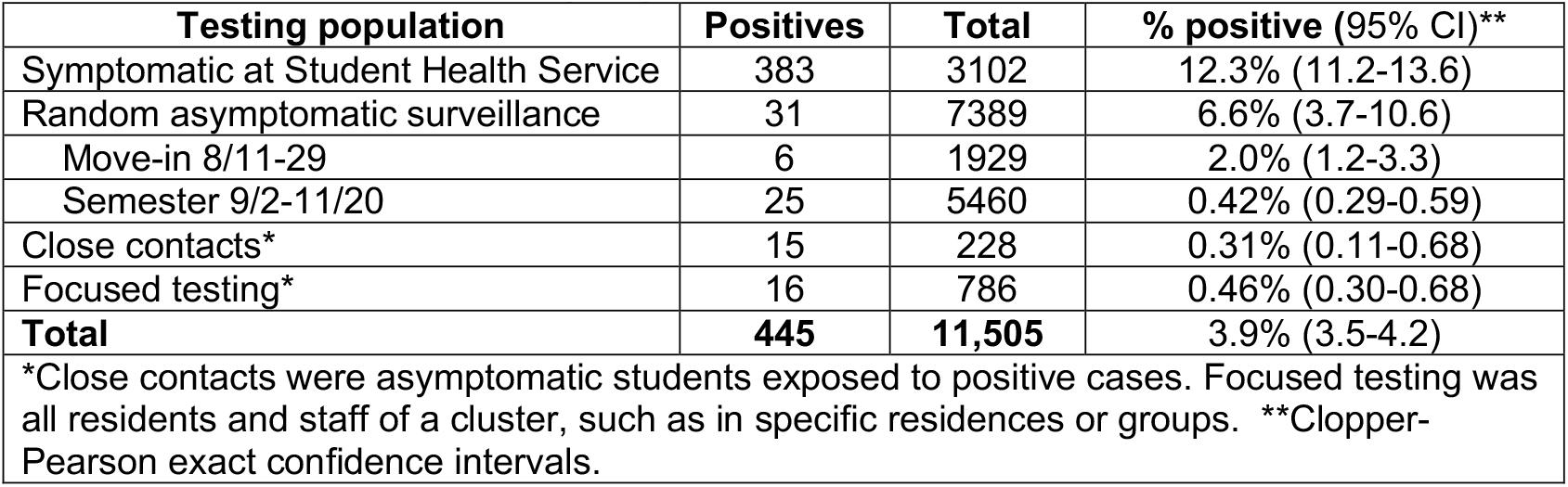
Results of student testing Aug. 11 to Nov. 20

| Testing population | Positives | Total | % positive (95% CI)** |
| --- | --- | --- | --- |
| Symptomatic at Student Health Service | 383 | 3102 | 12.3% (11.2-13.6) |
| Random asymptomatic surveillance | 31 | 7389 | 6.6% (3.7-10.6) |
| Move-in 8/11-29 | 6 | 1929 | 2.0% (1.2-3.3) |
| Semester 9/2-11/20 | 25 | 5460 | 0.42% (0.29-0.59) |
| Close contacts* | 15 | 228 | 0.31% (0.11-0.68) |
| Focused testing* | 16 | 786 | 0.46% (0.30-0.68) |
| **Total** | **445** | **11,505** | **3.9% (3.5-4.2)** |

\*Close contacts were asymptomatic students exposed to positive cases. Focused testing was all residents and staff of a cluster, such as in specific residences or groups. \*\*Clopper-Pearson exact confidence intervals.

**Figure 2.**
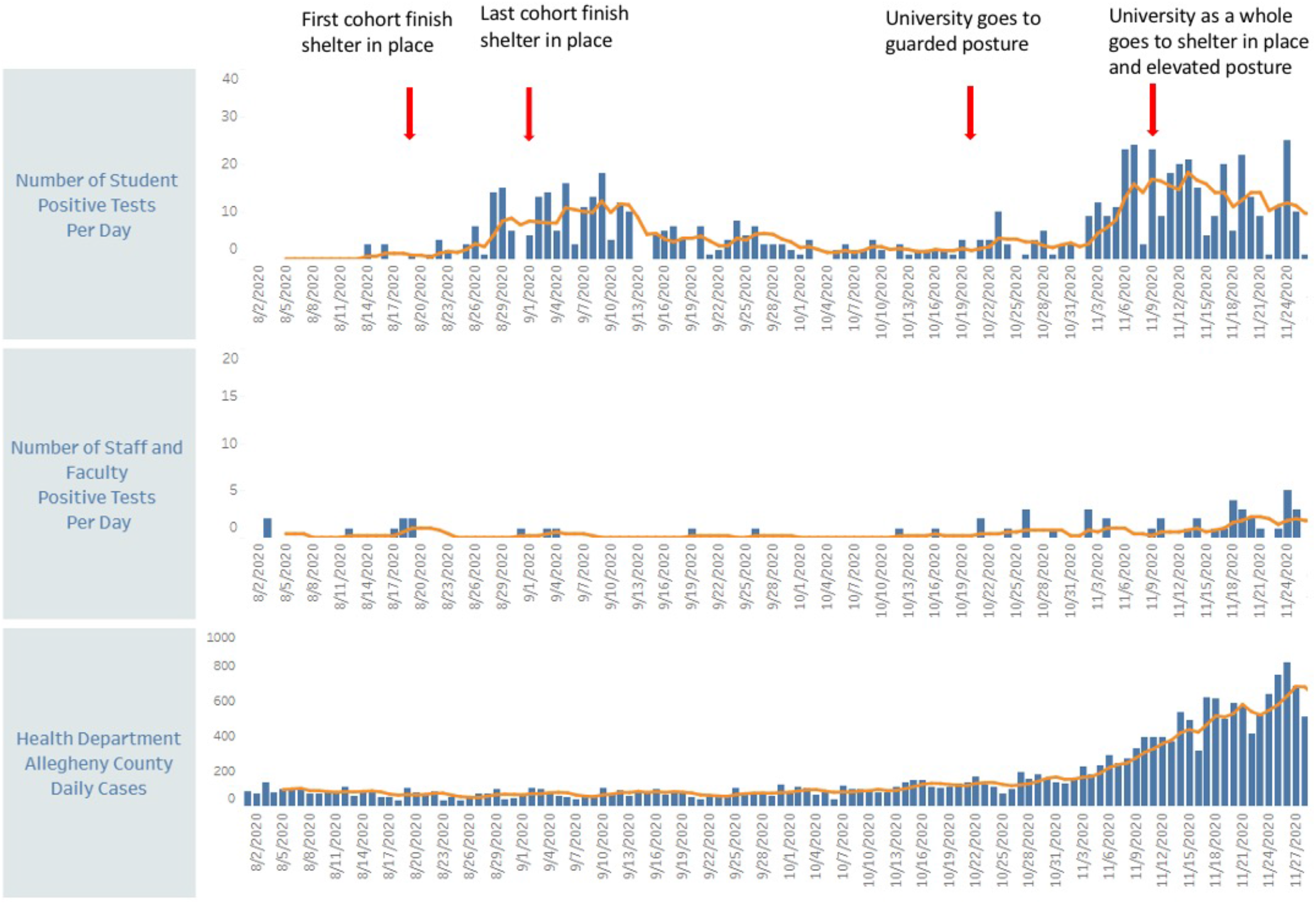
Number of SARS-CoV-2 positive individuals during the semester among students (top), staff/faculty (middle), and local county (bottom). Numbers above the graphs indicate 5-day rolling averages.

In November, cases increased exponentially in Allegheny County from ∼100 to >500 cases per day (**Figure 2**). At Pitt, the case count also increased, but the five-day rolling average did not exceed 20 cases per day(**Figure 2**). This surge was largely linked to off-campus socialization but did include some clusters in residence halls. The University again flattened the curve (**Figure 2**) via a communications campaign, increased focused and close contact testing, and targeted outreach to students emphasizing behavioral mitigation. The campus depopulation plan involved sheltering-in-place, which went into effect on Nov. 9, to help limit the spread of the virus. The number of student cases in isolation halved between Nov. 12 and 19. Staff and faculty cases rose due to community spread but remained relatively low with only 17 cases in isolation on Nov. 19. Throughout the term, the majority of positive cases occurred among undergraduates living off-campus (**Table 2**). Use of on-campus isolation beds peaked in November with 33.6% occupancy (97 of 289 isolation beds).

**Table 2.**
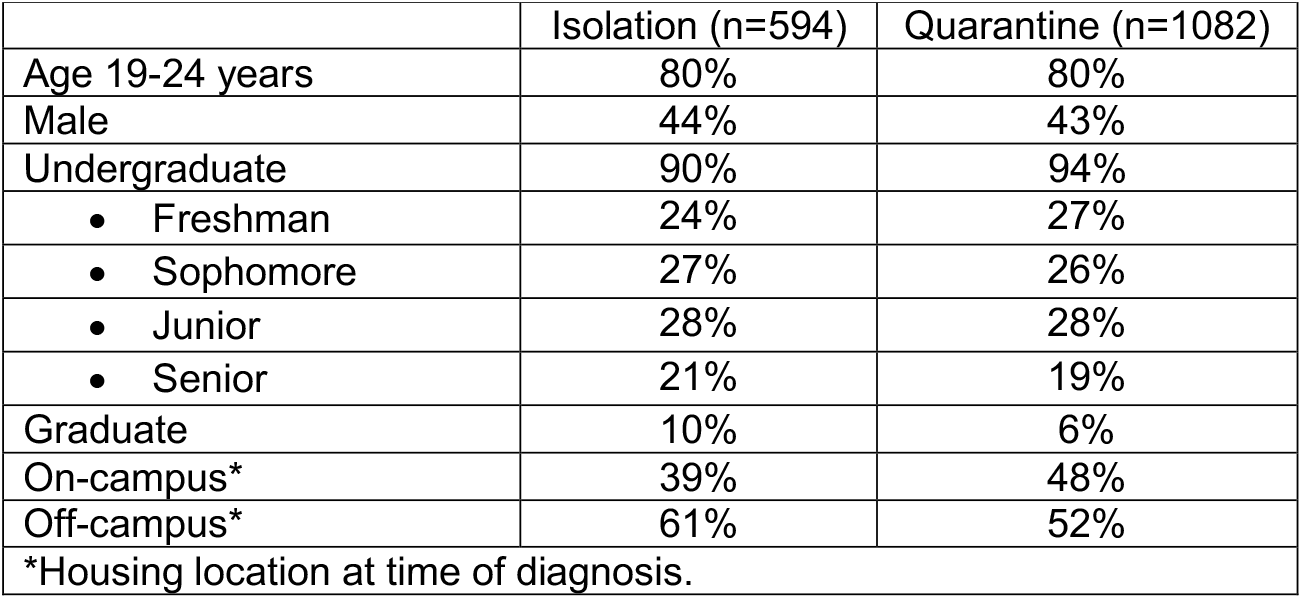
Demographics of Students in Isolation and Quarantine

|  | Isolation (n=594) | Quarantine (n=1082) |
| --- | --- | --- |
| Age 19-24 years | 80% | 80% |
| Male | 44% | 43% |
| Undergraduate | 90% | 94% |
| • Freshman | 24% | 27% |
| • Sophomore | 27% | 26% |
| • Junior | 28% | 28% |
| • Senior | 21% | 19% |
| Graduate | 10% | 6% |
| On-campus* | 39% | 48% |
| Off-campus* | 61% | 52% |

\*Housing location at time of diagnosis.

Seventy-four percent of students opened email testing invitations, and adherence to random asymptomatic testing by students who self-scheduled exceeded 90% during the semester. Of commercial testing offered free to all students after travel home, 6,170 kits were ordered and 3,720 samples were processed and reported, of which 77 (2%) were positive.

Housing data for on-campus student positive cases indicated that bathroom type—communal versus private— had no impact on the incidence of infection. This suggests that transmission did not occur in these shared spaces and that fomite transmission did not occur to any significant degree. Moreover, markedly different case rates occurred in similar residence environments (e.g., fraternities vs sororities). Contact tracing confirmed these findings, as clusters occurred in association with unsafe social gatherings (e.g., parties) or within shared residences that did not observe mitigation behaviors. Collectively, these data suggest that behavior—and not the physical housing arrangement—is abetting transmission.

Throughout the term, contact tracing identified multiple potential clusters to target with focused testing and additional education on COVID-19 mitigation strategies. Among all students testing positive, 29% were first identified as a close contact and already in quarantine at the time of diagnosis.

CMRO update emails served as a critical and direct line to students. Among students surveyed, 88% felt they received timely updates about COVID-19 and 77% considered the CMRO messages to be their primary source of COVID-19 information. These messages maintained strong opening rates of 48-63% (mean 53%) during the term. The “Casey” graphic (**Fig. 1**) reached >640,000 unique individuals on the University Facebook page and >27,000 on the University Twitter. This graphic was also shared widely, including by health departments, medical professionals and other universities.

From Aug. 24 to Dec. 4, industrial engineering students collected >13,000 observations of nearly 35,000 people from >60 locations around campus. Among those observed, approximately 78% were properly wearing a mask and about 72% were properly wearing a mask while also practicing physical distancing. This information was used to shape communication efforts and, near the end of the term, a 10-day moving average showed compliance levels >90%.

## DISCUSSION

We sought to limit viral transmission on a large urban university campus during the fall term. The primary tools were mitigation and communication, with testing serving as an important complementary component. Students were generally adherent with sheltering in place, masking, and physical distancing.

No classroom transmission occurred, and there was no evidence of transmission from students to faculty or staff. The majority of positive cases occurred among undergraduates—most were residing off-campus—and were related to unmasked social gatherings. There was no evidence of health sciences students (medical, dental, nursing, and allied health professions) becoming infected in, or transmitting within, clinical teaching environments. There were no student hospitalizations during the fall term despite >440 cases. In early November, the Pitt rate slowed and declined despite the surrounding county continuing to increase; this suggested that communications were altering student behavior. These results indicate that campus spread can remain contained despite exponential growth in the surrounding community.

Multiple platforms were necessary for effective communication coverage. Communication strategies were continuously revised to incorporate new knowledge, data, and feedback provided to better support both mitigation behaviors and students in isolation/quarantine. During each surge, these interventions were successful in affecting student behavior to flatten the curve. Flexibility was also key. The ability to quickly revert to a higher Risk Posture when necessary was effective. Virus control required adaptability, and rapid responses; videoconferencing enabled multiple stakeholders to meet quickly and easily.

The public data dashboard and CMRO’s twice-weekly email updates were well-received, though a balance was required between flooding the Pitt community with too much information and providing students with clear interpretations of the data. The message that testing helps identify infected individuals as a means of protecting others—not shortening quarantine—required frequent reinforcement. Students were also regularly reminded that a single negative asymptomatic test result did not preclude the possibility of recent exposure or the development of symptoms over the next few days. On the contrary, negative test results can provide a false sense of security that leads to reduced mitigation and increased spread, as documented in professional sports ^7, 8^, universities ^9-12^, and government organizations including the White House. Thus, students received repeated, regular messaging that a single negative asymptomatic test did not confirm whether someone has been recently exposed or if they would develop symptoms in the next few days. Students were reminded of this risk and encouraged to follow mitigation under the assumption that any person—themselves included—could be asymptomatically infected. Generally speaking, mitigation-related messaging sought to celebrate compliance successes and push out clear and concise behavioral nudges. As Pitt’s experience shows, this approach can be used to cultivate a shared sense of responsibility and engage and align members across a community in virus control.

However, behavioral messaging can be overdone. The IOC considered direct, more forceful messages. For example, even with a low asymptomatic prevalence of 0.4%, the Pitt community could be expected to have about 80-100 COVID-19 asymptomatic infections at any given time. Should students receive this information, and how would it be received? The IOC decided additional gain from such messaging would offer little encouragement over concrete mitigation messaging. Pitt’s experience suggests that careful behavioral nudges can promote a shared sense of community protection. We observed an increase in mask compliance during the term, and in a temporal fashion after communications about lower compliance. Moreover, during the surge in cases in early November, the Pitt case rate slowed and declined despite the surrounding county continuing to increase; this suggested that communications were altering student behavior.

Guidelines issued during Summer 2020 from the CDC and American College Health Association advised against universal testing of asymptomatic students, and updated recommendations from both agencies continue to note cautions regarding mass testing of asymptomatic students ^13, 14^. These cautions are due in part to the strain on testing resources and the adverse impact on mitigation behavior a negative test may provoke, due to the student’s belief they are “negative” and uninfected ^15^. Infected individuals typically test-negative during the first few days of infection, and yet may spread virus to cause outbreaks ^16-20^. Notably, the quality of virus control at Pitt was comparable to that of peer universities and colleges using universal testing strategies (**Table 3**). These schools were selected for comparison based on size, urban location, and publicly available dashboard data. Close contact testing and focused testing were effective in shutting down—and, ostensibly, limiting the development of—outbreaks and clusters. A recent analysis suggested that mitigation is a highly cost-effective non-pharmaceutical intervention ^21^.

**Table 3.** Comparison of Pitt student testing with anonymized peer urban institutions.

| School | Undergraduate enrollment | Dates | SARS-CoV-2 positive tests | Positive/10,000 | Tests performed | Tests/10,000 |
| --- | --- | --- | --- | --- | --- | --- |
| Pitt | 19,200 | 8/1 – 11/20 | 445 | 232 | 11,505 | 5,992 |
| School A | 6,700 | 8/2 – 11/20 | 152 | 227 | 158,817 | 237,040 |
| School B | 15,964 | 8/1 – 11/20 | 1,264 | 792 | 141,026 | 88,340 |
| School C | 18,000 | 7/27 – 12/7 | 572 | 318 | 467,000 | 259,444 |
| School D | 34,120 | 7/6 – 12/7 | 3,892 | 1,141 | 952,000 | 279,015 |
| School E | 40,640 | 8/7 – 12/7 | 4,353 | 1,071 | 83,458 | 20,536 |

Number of undergraduates, positive tests, and total tests obtained from public COVID-19 dashboards.

Our study has limitations. Notably, since we used a random subset surveillance strategy, the total number of SARS-CoV-2 infections at Pitt is an underestimate. In addition, some students sought testing at off-campus sites. However, we worked closely with the local health department, and many of these students’ results were reported to the Pitt contact tracing team and counted. Nonetheless, some cases may have been missed.

In summary, the three-pillar strategy of mitigation, communication, and containment—paired with the efficient and appropriate use of testing resources for asymptomatic students—produced effective virus control. Recent CDC guidance, as well as a report from the National Academies of Sciences, Engineering, and Medicine, emphasize the need for individualized approaches for IHE and a tiered approach to testing, with mass asymptomatic testing the lowest priority ^13, 22, 23^. It is worth noting that the goal of IHE SARS-CoV-2 mitigation programs is to support students’ health and well-being, facilitate the educational mission, and prevent uncontrolled outbreaks. We present here a model of successful virus control in a large, urban college setting using a targeted testing strategy as opposed to a universal mass testing strategy. Student involvement was the key to success in this model.

## Data Availability

Data is available on public dashboard

